# Measuring the growth of infectious disease modelling publications and their impact on policymaking: a Large Language Model-assisted bibliometric review

**DOI:** 10.1101/2025.06.12.25328864

**Authors:** Paula Christen, Malik H A Ahmed, Brandon Chua Wen Bing, Panupong Chaowanasawat, Emma Chapman-Banks, Yusuf Ozkan, Sabine van Elsland, Anne Cori, Sarin Sarin, Matthew D Whitaker, Marc Chadeau-Hyam, Saudamini Vishwanath Dabak, Mark Jit

**Affiliations:** Imperial College London; Imperial College; Health Intervention and Technology Assessment Program Foundation (HITAP); University College London; New York University

## Abstract

**Background:** Infectious disease modelling (IDM) is increasingly being used to understand disease transmission and inform public health policy. However, its growth and policy influence has never been quantified, possibly because of the volume of literature involved. The development of large language model (LLM)-assisted reviewing allowed us to quantify the expansion of IDM publications over time, trends in policy citations of IDM research, and regional disparities in research contributions and citations in policy documents.

**Methods:** An LLM-assisted bibliometric review was conducted using Embase, Medline, and Web of Science, identifying IDM publications from database inception to December 2024 using GPT-4o. Inclusion criteria encompassed peer-reviewed studies employing mathematical, statistical, or mechanistic models for infectious disease outcomes. LLM accuracy was iteratively refined by human review. We extracted publication metadata, geographic scope, and policy citations using Overton, a global database of policy documents. Growth trends were analysed using negative binomial regression models, and geographic disparities were assessed based on World Bank income classifications.

**Results:** A total of 33,255 IDM publications were identified over 44 years, with distinct growth phases. The LLM selection and data extraction achieved 98% and 100% accuracy respectively, compared to human search. Publication volume increased from the time of HIV/AIDS emergence, experiencing steady expansion through multiple outbreaks (Ebola, SARS, H1N1, MERS, Zika), and surged sharply just before the COVID-19 pandemic before declining post-2021. Recorded policy citations accounted for 1.7% of IDM publications, closely following the overall publication trend, peaking during periods of heightened public health attention. Policy citations largely reflected national research outputs, with notable cross-regional adoption of IDM evidence in some settings.

**Conclusion:** Strengthening the integration of IDM evidence into policymaking processes may require addressing geographic disparities in research output (and its recording in international databases), enhancing cross-regional collaboration, and improving mechanisms for policy uptake. Following the COVID-19 pandemic, policy citations declined despite continued growth in IDM literature, suggesting a potential lag or shift in policymaking priorities.

## 1. Introduction

Infectious disease modelling (IDM) has a long history as a critical tool for understanding disease transmission dynamics and informing public health policies and programs.^1,2^ IDM provides valuable insights into the population-level effects of interventions such as vaccination, antimicrobial stewardship, and non-pharmaceutical interventions. Moreover, it is an essential component of health economic evaluations of infectious disease interventions, guiding resource allocation decisions around disease control measures.^3^

IDM publications have grown in volume, but it is unclear if this is purely in line with the growth of publications in other scientific disciplines ^4,5^. It is also unclear if the growth in IDM literature has translated into greater influence on policy and program decision-making. Scientific developments such as ensemble approaches to synthesise multiple pieces of evidence to reduce decision-making uncertainty ^6^ have contributed to growth in the scientific literature. However, it is unclear whether such developments are valued by policymakers and public health officials, for whom societal, political and other factors may be more important than uncertainty characterisation or performance of predictions.^7,8^

Additionally, it is important to understand where published IDM research, since locally generated research is seen as more salient, credible and legitimate^9^, and thus more likely to influence local policymaking. Most scientific output is generated in high-income countries (HICs)^8,10^, with scientometric studies suggesting that this is driven by economic development^11,12^. IDM research may follow this pattern, since it requires access to computational resources, epidemiological data from surveillance systems and sustained research funding that is more prevalent in HICs. Additionally, authors from low- and middle-income countries (LMICs) may face language and cultural barriers, partly because journal offices, editors and reviewers are predominantly in HICs. Systematically tracking bibliometric trends in IDM literature has been difficult due to the large and growing volume of literature. However, recent advances in large language models (LLMs) have made this work more feasible. In this paper, we use LLMs to (1) quantify the expansion of IDM publications over time, (2) evaluate trends in policy citations of IDM research, and (3) analyse regional disparities in research outputs and their citations in policy documents.

## 2. Methods

### 2.1. Search strategy

We used an LLM-assisted approach to identify IDM research published on Medline, Embase and Web of Science. Our analysis involved five steps: (i) human-led database searching, (ii) LLM-assisted screening, (iii) LLM-assisted data extraction, (iv) data collection on policy impact, and (v) mapping of IDM literature and policy documents.

We included peer-reviewed publications published and indexed on Medline, Embase, Web of Science before 10 December 2024. We searched the databases using title, abstract, and keywords only. To make such an expansive scope possible, full texts were not retrieved, screened, or analysed, and grey literature (e.g., policy briefs, reports, theses, and unpublished studies) was not included. The search strategy was reviewed by a librarian. Search protocols used in each database are in S1. Embase and Medline searches were done using the Ovid platform.

### 2.2. Screening

Studies were included if they featured: (1) mathematical models, i.e. representations of a real-world system using numbers and/or equations. These include both statistical models (e.g., regression, machine learning) and mechanistic models (e.g., compartmental models, agent-based models, discrete-event simulations); (2) original primary peer-reviewed research published in scientific journals in English; (3) clinical, health systems, and/or economic outcomes of infectious diseases in humans; and (4) disease-linked conditions where the infectious disease was explicitly modelled. Studies were excluded if they: (1) lacked mathematical modelling; (2) presented no clinical, health systems, or economic outcomes; (3) focused exclusively on non-human outcomes or non-infectious diseases; (4) were not primary research; (5) were not journal publications; (6) were purely empirical, conceptual or descriptive studies without mathematical components; or (7) non-English language publications.

We developed and implemented a dual-stage screening framework integrating human review with LLM classification using GPT-4o^13^. Initially, the inclusion and exclusion criteria were supplied to the LLM alongside a prompt to parse the article abstract and determine inclusion or exclusion based on the criteria. A random sample of 111 papers screened by the LLM were compared against human decisions. Discrepancies were used to refine the prompt to the LLM. This iterative process continued until the automated classification achieved 98% accuracy against human review (n=111 randomly selected records). S2 provides further details on the methodology. S3 Figure 1 shows an overview of the study selection process.

### 2.3. Data extraction

To identify the geographical scope of included studies, we developed an automated extraction system using a large language model (GPT-4o) to process titles and abstracts. Through multiple iterations of human review and refinement (S3, Figure 1), the prompt (S3) was refined to identify and standardise location information according to specific rules: studies were labelled as “Global” only when explicitly stated with no specific locations mentioned; regional or country-specific when locations were explicitly stated; and “NA” when no location information was provided in either the title or abstract. For studies spanning multiple sites, each location was recorded with standardised semicolon separation (e.g., “Brazil; Portugal; France”).

When subnational-level information was available, it was recorded with its corresponding country (e.g., “Lausanne, Switzerland”). The pipeline’s accuracy was validated through human review of 100 randomly selected records, achieving 100% accuracy, and enabling reliable analysis of the geographic distribution of IDM research.

### 2.4. Data collection on policy impact

Policy citations were identified using the database Overton (see overton.io).^14^ This database includes over 12 million policy documents from governments, official bodies, intergovernmental organisations (IGOs), and think tanks from nearly 200 countries, linked to the evidence they cited. Using an Overton API key, for each peer-reviewed publication, the number of citations in policy documents, documents citing the IDM publication, the policy document title, the date of policy document publication, and its type of organization as categorized by Overton (government agencies, intergovernmental organisations, think tanks, and other organisations) and country were extracted.

### 2.5. Data analysis

To investigate trends in IDM research and its influence on policy, we compared publication volume and policy citations with the timing of major infectious disease outbreaks. The timeline of these outbreaks was extracted from the Council on Foreign Relations’ record of significant epidemics in the modern era (https://www.cfr.org/timeline/major-epidemics-modern-era).

To estimate the growth rate of IDM publications over time, we applied segmented negative binomial regression models (see S5 for details). Models with one, two, and three breakpoints were compared, and the final model was identified using the Akaike Information Criterion (AIC). The doubling time of IDM publications was computed using the exponential coefficient of the respective time period, calculated as ln(2)/growth rate.

We quantified the proportion of IDM publications cited in policy documents and analysed their geographic research scope distribution based on country income classification following the World Bank 2019 income rankings (Organisation for Economic Co-operation and Development (OECD) high-income, high-income non-OECD, upper-middle-income, lower-middle-income, and low-income countries). The relative frequency of citations from each income group was assessed, along with the ratio of foreign to national publication citations in policy documents for each country. We generated geographical maps to visualise the global distribution of IDM literature (by country focus) and the aforementioned ratio. Additionally, we evaluated trends in IDM publication volume across income classifications to assess whether patterns of research output and policy influence varied by economic status.

All source code used for data analysis, which was performed in R, is available on GitHub (https://github.com/paulachristen/idm_literature)^15^.

## 3. Results

### 3.1. Volume and Trends in IDM Publications

A total of 33,255 IDM publications published over 44 years were identified (Figure 1). The most parsimonious model segmented growth into four phases, with breakpoints in 1989, 2018, and 2021 (S6, Figure 2).

**Figure 1.**
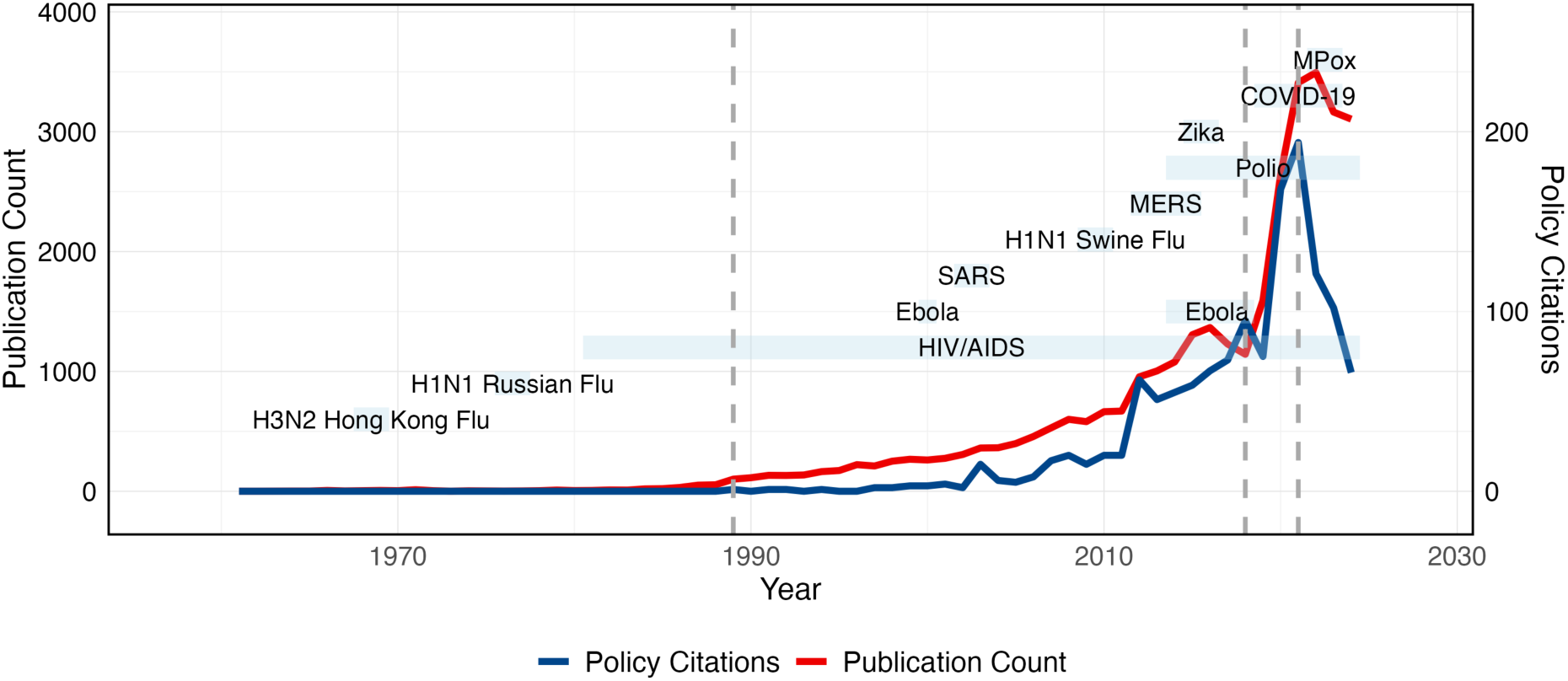
Timeline of major epidemics, infectious disease modelling publications identified and citations of the infectious disease modelling publications in policy documents. Vertical, grey dashed lines indicate the sections of the segmented negative binomial regression models, with the number of breakpoints determined using the Akaike Information Criterion (AIC).

**Figure 2.**
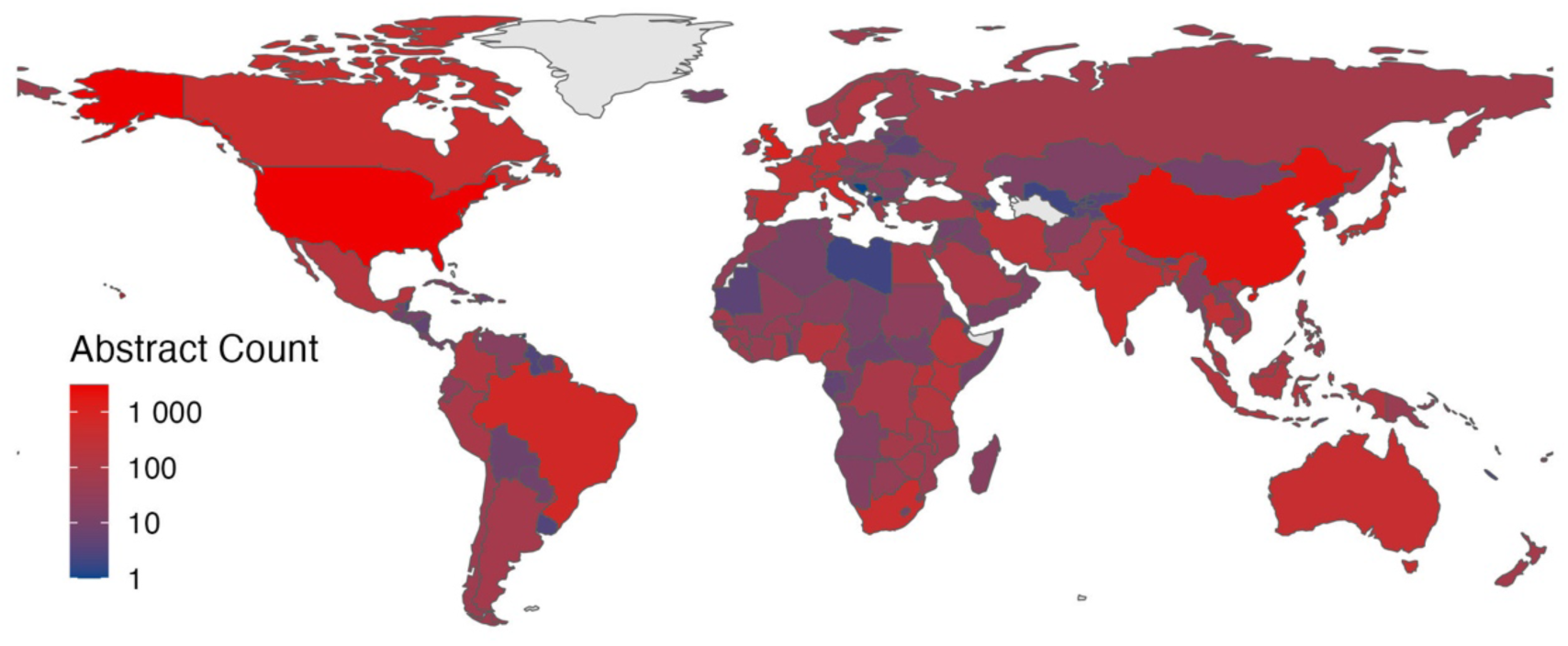
Global distribution of geography-specific infectious disease modelling literature published between 1980 and 2024 identified in three literature databases (Medline, Embase, Scopus) on log scale.

From 1980 to 1989, the exponential growth rate was approximately 0.3275 per year, resulting in a doubling time of 2.1 years. From 1989 to 2018, the growth rate slowed to 0.0917 per year, with a doubling time of 7.6 years. Between 2018 and 2021, the growth rate surged to 0.3640 per year, with a doubling time of 1.9 years. This period ended just before the COVID-19 pandemic. Between 2018 and 2021, the growth rate increased to 0.3640 per year, with a doubling time of 1.9 years. After 2021, mid COVID-19 pandemic, a decline in publication activity is noted with a negative growth rate (-0.0598 per year). 60% of IDM publications specified a geographic scope in their abstract. For 7% of these publications, no countries could be extracted from the abstract as these covered regions or multiple countries (e.g., 13 European countries, sub-Saharan Africa, or the Amazonas region). Less than two percent (1.9%) of the literature has a global scope; 47.8% of the literature focuses on Europe and the Americas together (S7, Figure 3). Country-specific IDM literature mostly focuses on the US and China (Figure 2).

### 3.2. Policy Citations of identified IDM Publications

Of all publications identified, 1.7% were cited in policy documents on the Overton database. Government agencies such as the Centres for Disease Control and Prevention (CDC) were identified as the most frequent citers (Figure 3). No policy documents citing IDM literature were found in the database for 74% of countries globally, mostly high-income (non-OECD), upper-middle-income, lower-middle-income, and low-income countries. Just like trends in IDM literature itself, growth in policy documents citing IDM literature reversed after 2021.

**Figure 3.**
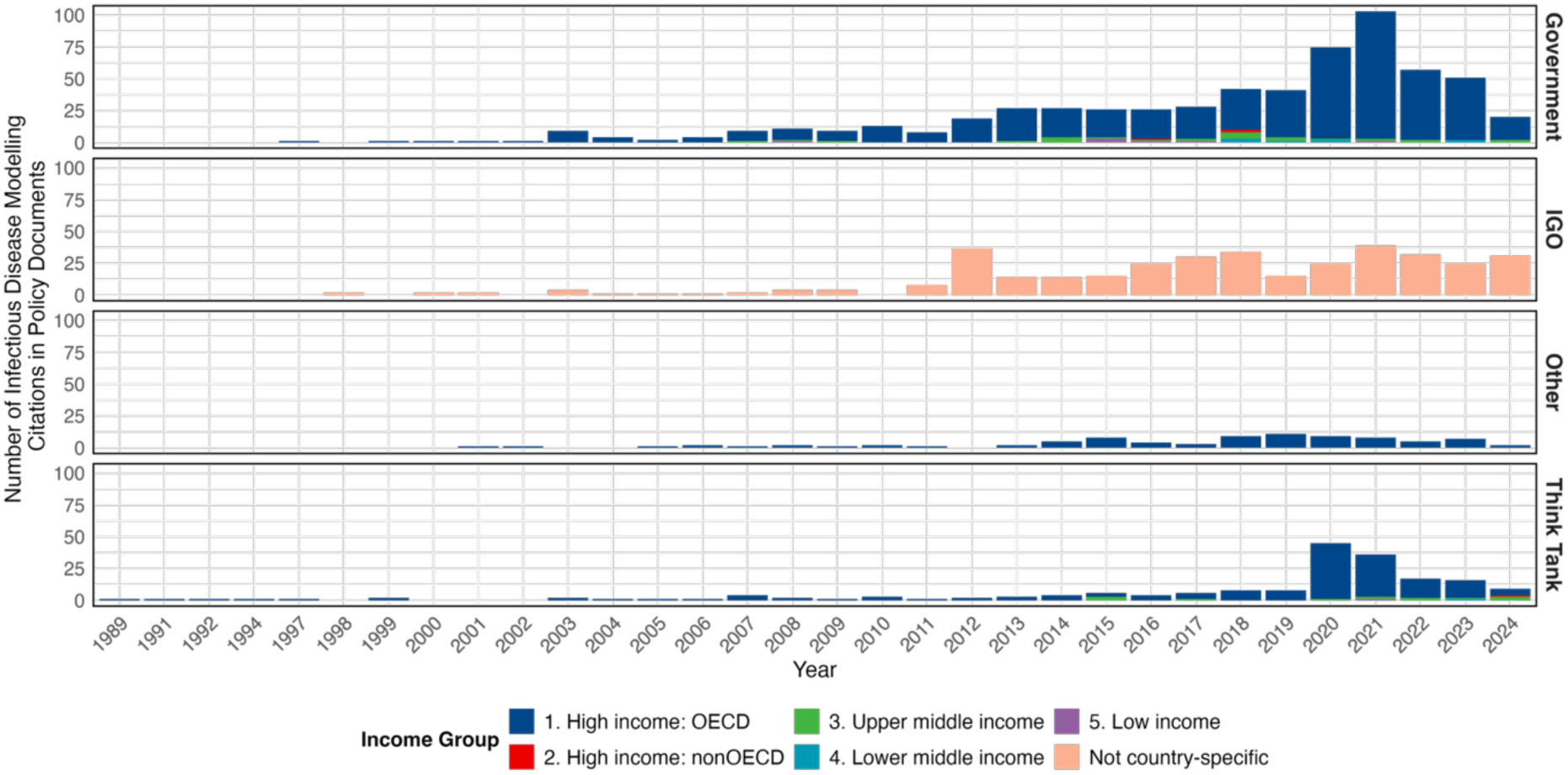
Number of citations of infectious disease modelling publications in policy documents identified in the Overton database over time, by type of organization and country income group. See S8, Table 3 for high-income OECD countries citing infectious disease modelling publications.

Of those that cited geography-specific IDM literature in their policy documents, central European countries cited most foreign IDM literature compared to national IDM literature. Among countries on which Overton has publications on, the US and the Philippines cited almost as many national as foreign IDM publications and none of the countries cited more national than foreign IDM publications in policy documents identified. Further, IDM literature was mostly cited in government policy documents (52%), followed by intergovernmental organisations (28%), think tanks (14%), and other organisations (6%). 25% of policy documents published by government entities were healthcare agencies (Table 1). Think tanks only started citing IDM publications from 2020 onward, while governments and international organisations increasingly cited more IDM publications over time (Figure 3).

**Table 1.** Citations of IDM publications identified in the Overton database by type of organization.

| Type | Sub-type | Number of policy documents citing IDM literature | Percentage |
| --- | --- | --- | --- |
| Government | Not specified | 377 | 28% |
|  | Healthcare agency | 177 | 13% |
|  | Agency | 76 | 6% |
|  | Research centre | 22 | 2% |
|  | Food and drug safety | 21 | 2% |
|  | Bank | 7 | 1% |
|  | Legislative research | 7 | 1% |
|  | City | 5 | 0% |
|  | Academy | 3 | 0% |
|  | Technology assessment | 1 | 0% |
| Inter-governmental Organisations | Healthcare agency | 212 | 16% |
|  | Not specified | 92 | 7% |
|  | Development bank | 62 | 5% |
|  | Research centre | 2 | 0% |
| Think tank | Not specified | 139 | 10% |
|  | Research centre | 30 | 2% |
|  | University affiliated | 14 | 1% |
|  | Consultancy | 4 | 0% |
|  | Industry association | 1 | 0% |
| Other | Aggregator (e.g., Cochrane reviews, clinical guidelines by independent review committees) | 85 | 6% |

## 4. Discussion

### Key findings

IDM literature has expanded over most of the time period examined (1989–2024). While other scientific literature has also grown, IDM literature experienced significantly faster growth. Bornmann *et al*. ^4^ estimated a growth rate for published scientific literature across four databases of 4.10% per year between 1670 and 2018 with a doubling time of 17.3 years, whereas IDM literature has expanded much faster with a doubling time of 2.1-7.6 years (depending on the period). The literature expanded rapidly in the 1980s (around the time of HIV/AIDS emergence), but slowed down after that, while still expanding faster than general scientific literature, before accelerating in the late 2010s. Policy citations of IDM research mirrored this trajectory, also exhibiting exponential growth. Notably, both IDM publications and policy citations peaked at the same time, suggesting close alignment in the timing of scientific production and its uptake in policy documents, although our annual-scale analysis may obscure shorter-term lags. However, this growth was not sustained in the aftermath of the COVID-19 pandemic.

We found geographical disparities in the identified IDM publications, with few publications in South Asia and Africa, despite these regions bearing a disproportionately high burden of infectious disease morbidity. Our findings concur with a systematic review that found that during the COVID-19 pandemic, most IDM research in Africa was conducted in just three countries^16^, highlighting systemic imbalances in capacity. This may be attributed both to an inherent bias in the literature databases (see below) as well as to the concentration of modelling capacity in high-income countries, limited funding as well as competing priorities in other countries.

### Limitations

The research has several limitations. Firstly, we limited the search strategy to English-language publications by applying an explicit language filter during database queries to ensure consistency in analysis and interpretation by the LLM. This approach, although estimated to cover 95% of relevant literature^17^, may in particular exclude many non-English language papers published in LMICs. Additionally, grey literature not formally indexed in academic databases (e.g., preprints, government reports) was excluded unless cited via Overton, which may indirectly capture such references through citations in indexed IDM literature.

The analysis is also constrained by the scope and coverage of Overton, which indexes public policy documents but does not encompass all policy documents ever written. Overton’s coverage is more comprehensive for policy organisations in HICs, with developed countries having stronger digital footprints being better represented. Additionally, 80% of the policy documents indexed in Overton were published after 2012, reflecting both the growth in policy documentation and digitisation practices during this period. Citation data from Overton also has limitations, as it targets a minimum accuracy of 98% and recall of 80% for scholarly documents, but issues may arise with scholarly papers not indexed by Crossref or those belonging to a series.

The choice of databases – Medline, Embase, and Scopus – was made to improve coverage and recall, but it is recognised that not all relevant literature may be captured. Medline, produced by the US National Library of Medicine, covers international biomedical literature, while Embase includes abstracts from biomedical and pharmaceutical journals. Scopus, a multidisciplinary tool, was chosen to capture models that may not be in conventional medical library outlets. Despite these efforts, reliance on these databases may still result in gaps in the literature review, particularly due to the exclusion of publications from Chinese-language journals in regional databases such as Wanfang and CNKI, which were not included in this search. The databases’ poor coverage of journals based in LMICs and Chinese-language journals may again have led to underestimation of the global literature.

Some abstracts did not specify all countries when the scope was broad (instead using terms like “sub-Saharan Africa” or “15 European countries,”) which may limit the granularity of geographical analysis, since data extraction was limited to titles and abstracts. Additionally, whilst our preliminary results on a base-truth sample of 111-human-reviewed title/abstracts showed LLM accuracy that exceeded 98% (109/111) in each iteration with zero false-negatives with our final prompt (S4), this is small compared to the 492,804-publications dataset screened. LLMs used for screening and information extraction may be subject to error and hallucination, although previous research suggests that screening by LLMs is more accurate than screening done by humans.^18^

### Recommendations for research and policy

Further research is needed to capture and index literature and policy documents produced in LMICs and not in English, so that the disparities in the use of IDM evidence for policymaking can be clearer.

Our bibliometric analysis did not distinguish between original research and more derivative works. Future research could identify countries producing the most innovative research using novelty scores, such as those developed by Shi and Evans^19^, and implemented by DeSci^20^ to quantify innovation by analysing semantic linkages between keywords and reference patterns in manuscripts. The algorithm compares manuscripts to historical publications and forecasts future trends, measuring a study’s novelty as its divergence from projected trajectories. This approach shifts focus from research volume to originality, offering a more nuanced perspective on how scientific advances influence policy. However, such an evaluation might undervalue work that is not scientifically “novel” but may still be useful for policy. This is particularly relevant in LMICs, where resource constraints may limit blue-sky discovery science in favour of work done rapidly to directly answer a policy question.

A systematic analysis of policy documents could offer deeper insights into the role of IDM research in decision-making.^21^ While previous studies have explored the use of modelling evidence in specific contexts - such as intervention spaces (e.g., vaccines^22,23^), stakeholder groups (e.g., World Health Organization guidelines^24^), or specific outbreaks (e.g., COVID-19^25,26^, H1N1 influenza^27^) – a broader, cross-sectoral approach could enable a more comprehensive comparison across time and various public health domains.

This study also demonstrates the potential for LLMs to support interrogation of the academic literature at scale. Manually screening 492,804 papers would require 16,427 hours (1.9 years) of human review at a rate of one abstract screened every two minutes. The LLM-assisted screening took less than three days, and had high accuracy compared to human reviewers.

## 5. Conclusion

This study quantified the rapid growth of infectious disease modelling (IDM) research, bookended by the global health crises of HIV/AIDS and COVID-19. It also revealed gaps in its translation into policymaking. Geographic disparities appear to persist, which could reflect both lack of institutional capacity in LMICs, although language and database limitations preclude definitive conclusions about this. Post-pandemic declines in both literature and policy citations suggest shifting priorities in evidence generation and uptake. To bridge these gaps, fostering equitable research collaboration, prioritising context-specific modelling in high-burden regions, and strengthening mechanisms for policy engagement are critical.

## Data Availability

All data produced are available online at https://github.com/paulachristen/idm_literature.

https://github.com/paulachristen/idm_literature

## Acknowledgements

We thank Nathalie Corneé and Kate Holvey for valuable insights on the Overton platform. This study was conducted as part of the Lancet Commission for Strengthening the Use of Epidemiological Modelling of Emerging and Pandemic Infectious Diseases.

## Supplement

### S1 Search protocol

#### Web of Science

(TS=(infect* OR “communicable disease” NOT “non-communicable disease” OR “disease transmission” OR pathogen OR transmit* OR zoonos* OR vector-borne OR bacteri* OR virus * OR viral* OR fung* OR protozoan OR helminth* OR prion* OR parasite OR “disease outbreak” OR epidemic OR “endemic disease” OR endemic)) AND TS=(“Infectious disease model*” OR “mathematical model*” OR “statistical model*” OR “computational model*” OR “transmission model*” OR “forecast*” OR “nowcast*” OR “epidemiological model*” OR “economic model*” NOT “within-host model*”)

#### Ovid – Medline® / PubMed

**Table 1.**
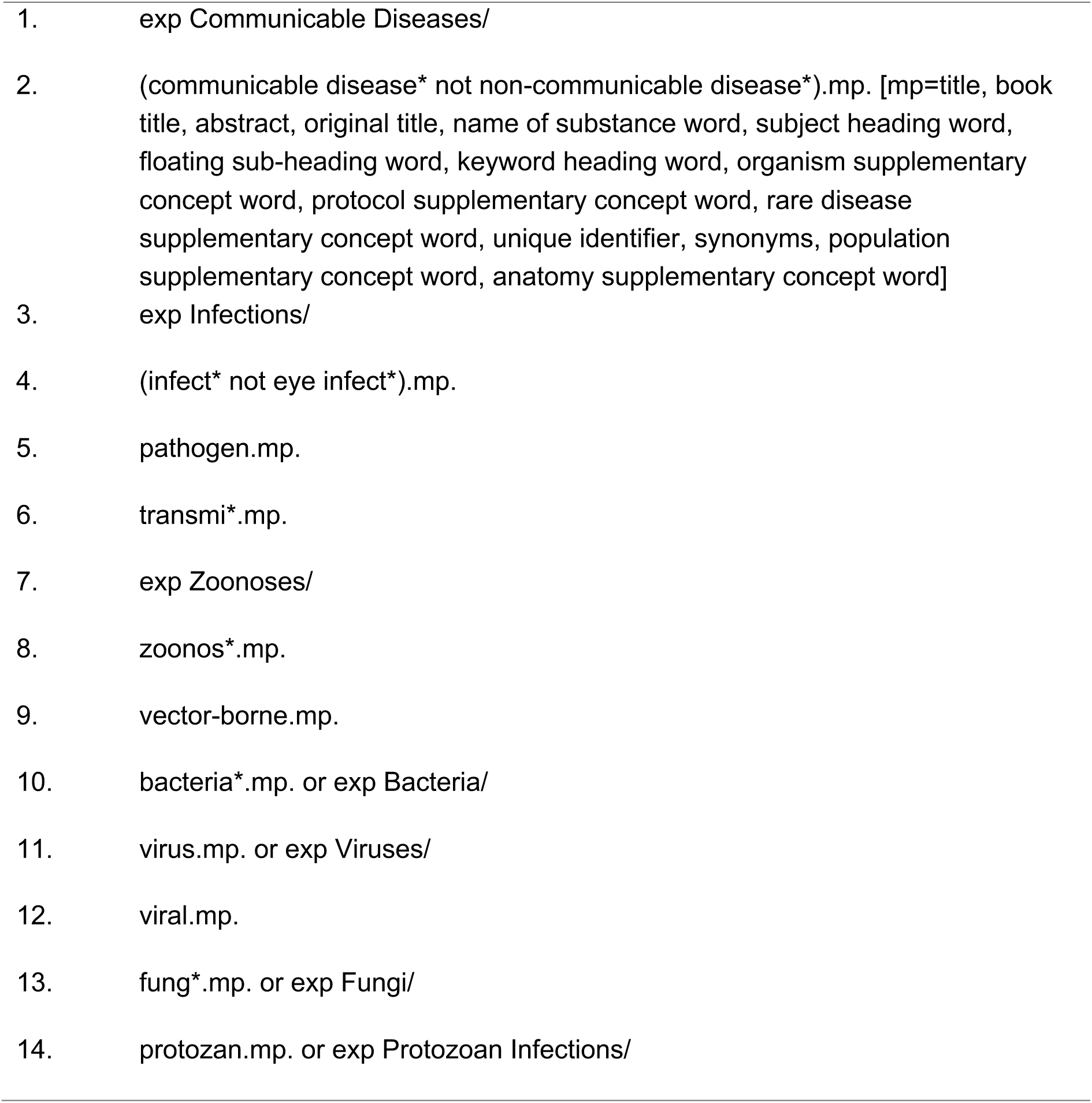
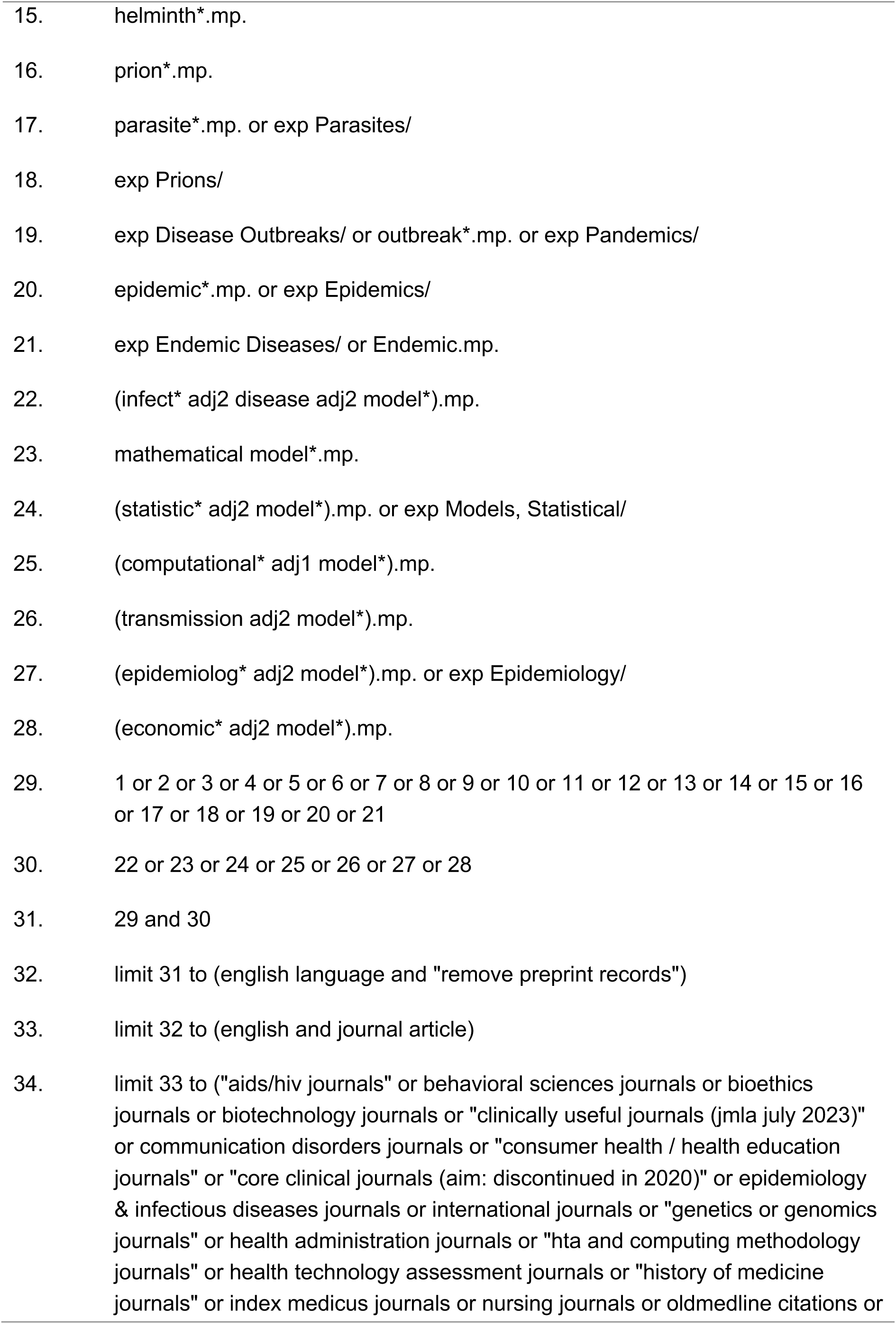
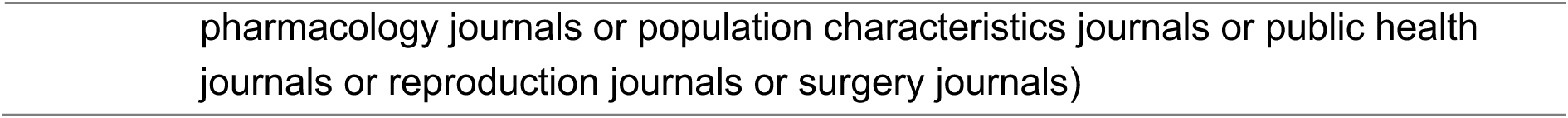
Medline® / PubMed search protocol.

#### Ovid – Embase

**Table 2.**
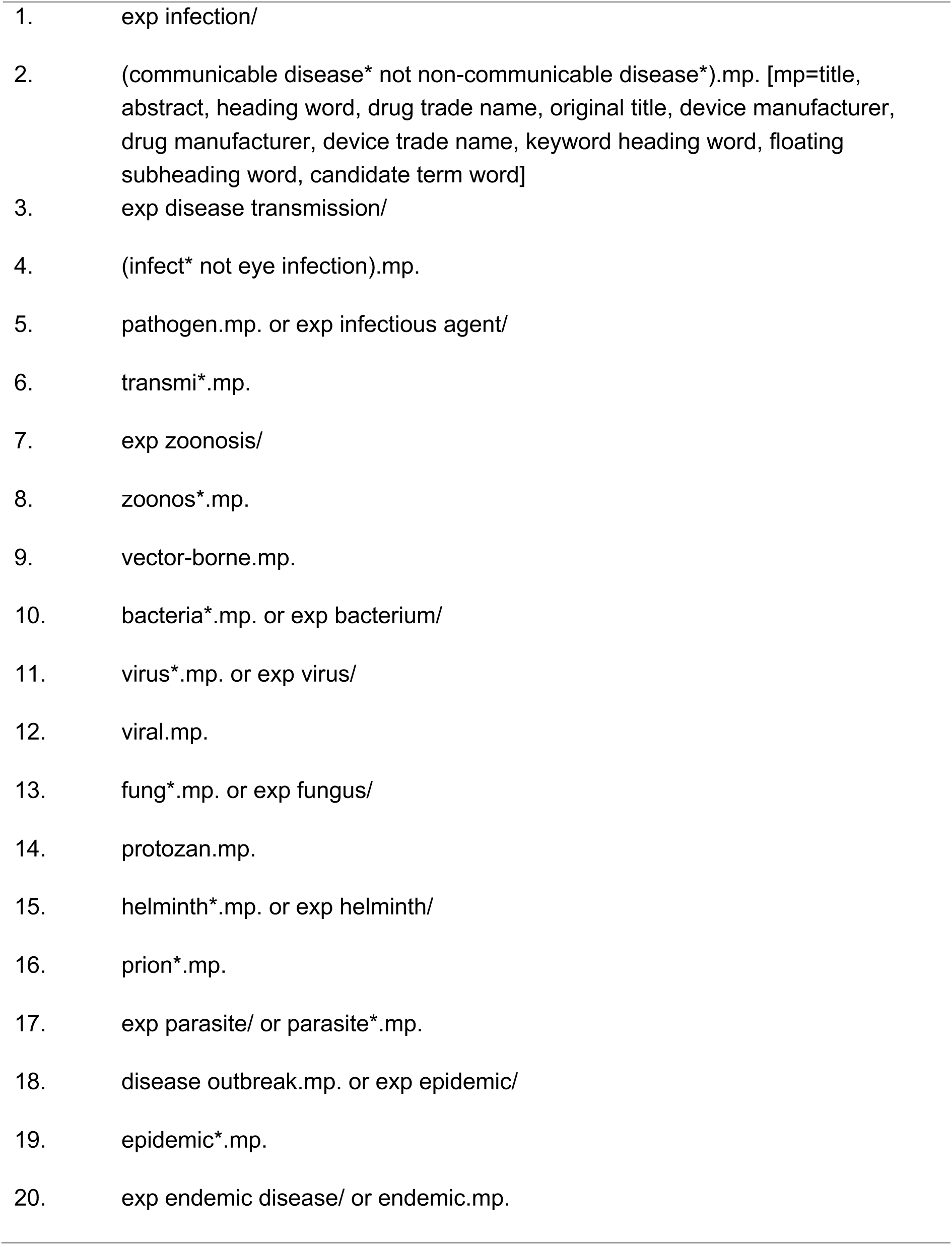
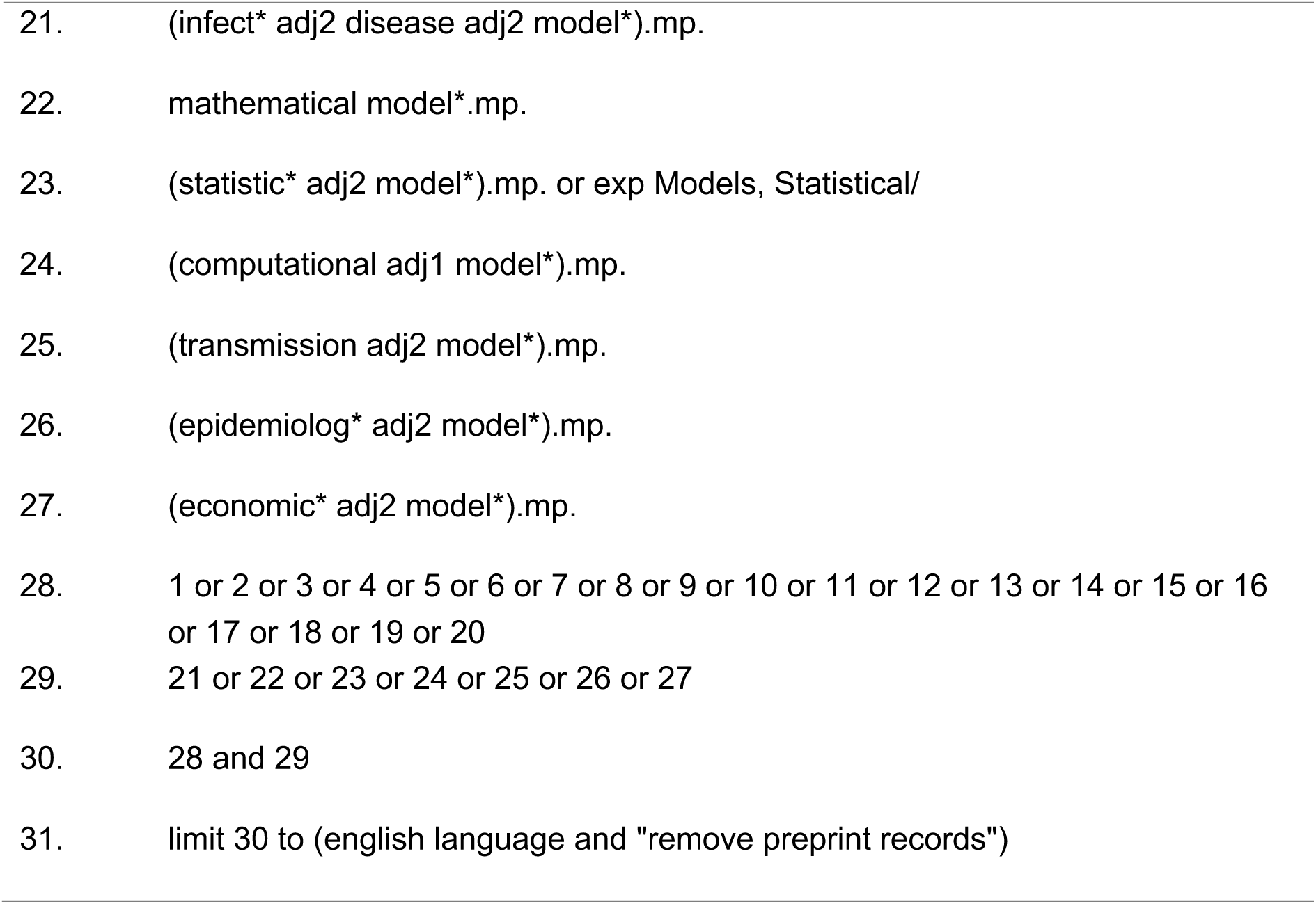
Embase search protocol.

### S2 Methodology

We implemented mostly standard rapid review method to retrieve and screen the literature, followed by the extraction of policy-related metadata and geographical information from the included papers for quantitative synthesis. The main difference was the inclusion of LLM assistance in the process.

#### 1. Literature Retrieval from Search Databases

The process began with a comprehensive literature search across multiple databases, Medline, Embase, and Web of Science, to identify relevant records. A search strategy was developed in collaboration with a librarian and a domain expert to ensure thorough coverage of the topic, incorporating appropriate keywords, Boolean operators, and database-specific filters. Given the potential for an extensive number of search results—exceeding 100,000 records—we anticipated challenges related to data download limits imposed by some databases. To address this, we adapted our retrieval strategy, such as segmenting the search into manageable portions and running the search strategy on multiple search database accounts simultaneously, to successfully compile the full dataset.

#### 2. Human-Informed Screening on a Subset

Following retrieval, we selected a subset of the literature for manual screening to establish a baseline for subsequent automated processes. This subset was carefully curated to include an equal number of papers that met our inclusion criteria and those that did not (exclusions), as determined by reviewers. The balanced composition of this subset minimized bias and provided a representative sample for evaluating the screening process. Three human experts reviewed each paper in the subset, applying predefined inclusion and exclusion criteria, and their decisions served as the ground truth for the next step. To ensure all reviewers had a shared understanding of inclusion and extraction criteria, 30 abstracts were double-screened and discrepancies discussed.

#### 3. Trial Prompt on the Subset to Evaluate Accuracy

Using the same subset from the human-informed screening, we trialled an automated screening prompt (see S4) to assess its accuracy in replicating the human decisions. The prompt was applied to the subset, and its performance was evaluated by comparing its outputs (inclusions and exclusions) to the human-established ground truth. Accuracy metrics were calculated to quantify the prompt’s effectiveness in correctly identifying relevant papers.

#### 4. Iterative Improvement of the Screening Prompt

If the trial prompt’s accuracy did not meet our predefined thresholds (e.g., sufficiently high sensitivity and specificity), we refined it by revising the criteria or incorporating additional features. The updated prompt was then reapplied to the same subset, and its accuracy was re-evaluated. This iterative process continued until the screening prompt achieved a satisfactory level of performance (we determined this to be 98% accuracy; other projects may have different tolerances for human-LLM discrepancy), ensuring reliability when scaled to the full dataset.

#### 5. Screening of All Papers

Once the screening prompt was satisfactorily refined, we applied it to the entire dataset of retrieved papers. This step leveraged the optimized automated process to efficiently filter the large volume of literature, retaining only those papers that met the inclusion criteria.

#### 6. Retrieval of Digital Object Identifiers (DOIs)

From the papers included after screening, we extracted their Digital Object Identifiers (DOIs). This step was critical for linking the screened papers to external resources in the subsequent phase of the methodology.

#### 7. Integration with Overton via API

The retrieved DOIs were submitted to the Overton platform using its Application Programming Interface (API). Overton is a database that tracks citations of academic papers in policy documents, providing metadata on how the literature influences policy contexts. By feeding the DOIs into Overton, we obtained detailed metadata about policy documents citing our included papers, offering insights into their broader impact and relevance beyond academic research.

#### 8. Extraction of Geographical Locations from Abstracts

Finally, we analyzed the abstracts of the included papers to extract geographical location information. This process mirrored the screening step in its systematic approach to identify and categorize location-based entities mentioned in the text. The extracted location data enriched our dataset, enabling analyses of regional trends or the geographical focus of the literature.

### S3 Modified PRISMA flowchart of the LLM-assisted study selection process

**Figure 1.**
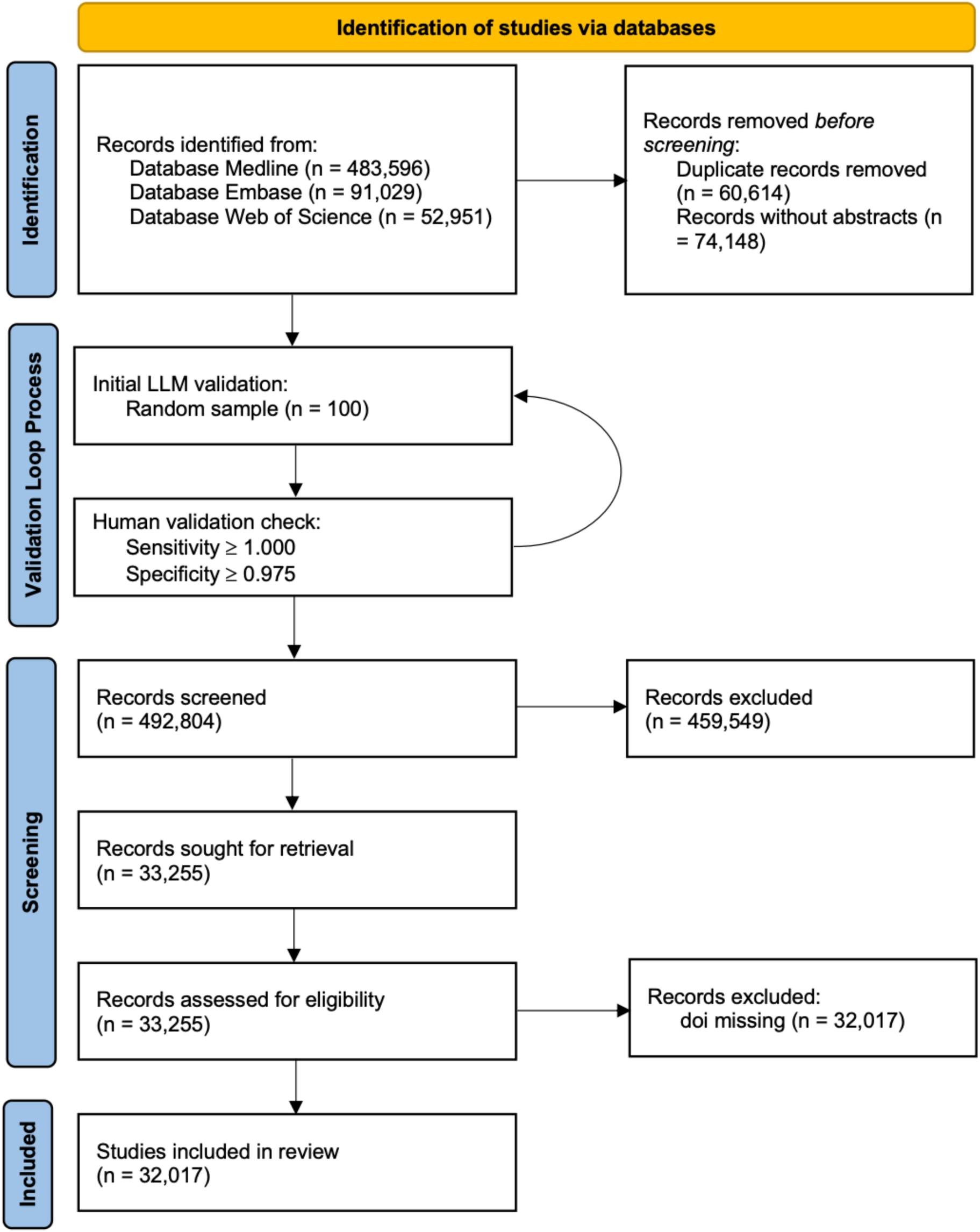
Modified PRISMA flowchart of the LLM-assisted study selection process.

### S4 Prompt fed to Chat-GPT 4.o

system_prompt = """Provide comprehensive and detailed responses to user inquiries. Ensure accuracy and completeness in information. Verify facts when necessary. Maintain a neutral tone, and avoid personal opinions or biases. Be concise. For abstract screening, the focus of the abstract must be on infectious disease."""

user_prompt = f"""

Given the following abstract, determine whether it meets the following inclusion or exclusion criteria for a literature review. Do not negotiate the criteria:

INCLUSION CRITERIA:

* Mathematical model, including statistical models (such as regression models and machine learning models), mechanistic models (such as compartmental models, agent-based models and discrete-event simulations) and sensitivity analyses;

* Original primary peer-reviewed research published in a scientific journal;

* Presents at least one clinical (in humans), health systems and/or economic outcome of infectious diseases;

* Behaviour / attitude is not considered a relevant health outcome;

* Health outcomes within an infected population (e.g., outcomes within HIV patients);

* Affects post infection (e.g., post-polio) or linked conditions (e.g., liver cancer is linked to hepatitis infection, or cervical cancer, linked to HPV) only if the linked infectious disease is modelled;

* Rheumatic heart disease if bacteria by which it is caused is modelled;

* Antibiotic prophylaxis to prevent surgical site infections or surgical site and nosocomial infections;

* Estimates of infectious disease modelling parameters/metrics (e.g., reproduction number, next generation metrix, etc.) as long as it is applied to an infectious disease (e.g., infectious disease mentioned, or direct link to infectious diseases at large is made)

EXCLUSION CRITERIA:

* Not presenting a mathematical model;

* Model without clinical, heath systems and/or economic outcomes e.g. within-host models, environmental models;

* Studies without a human health outcome e.g. models of plant, non-human animals, microorganisms or environmental outcomes;

* Studies without primary research e.g. systematic reviews and meta analyses, protocols;

* Studies not published as full research articles in journals e.g. conference proceedings, abstracts, posters;

* Empirical (wet lab, clinical or field-based) study (including trials, clinical reports, surveys and interviews) without any modelling component

* Behaviour / attitude / awareness as outcome;

* Social media usage; information seeking behaviour;

* Reporting descriptive statistics only;

* Non-clinical mental health outcomes;

* Vector transmission risk;

* Phylogenetic studies;

* Purely theoretical

Finally, classify the abstract as “Include” or “Exclude” based on the criteria, and provide a brief explanation for your decision.

Index:

{index} Title:

{title} Abstract:

{abstract}

Output the decision in the following JSON format:

{{

        “index”: “{index}”,

        “title”: “{title}”,

        “abstract”: “{abstract}”, “decision”: “Include” or “Exclude”,

        “explanation”: “<BRIEF explanation>”

}}

"""

### S5 Regression model

#### Base Poisson Model

The basic model is:

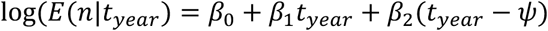

where:

- *n* is the count response variable;
- *t_year>_* is the predictor (time);
- *β*_0_ *and β*_1_ are the intercept and slope respectively.

#### Segmented Models

1. One-Segment Model

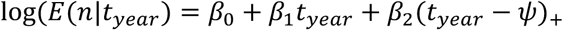

where:

- *Ψ* is the estimated breakpoint.
- (*t_year_* − = *max* {0,*t_year>_* − *Ψ*}  is the positive part function, which equals zero when *t_year_* ≤ *Ψ* and *t_year_* − *Ψ* when *t_year_* > *Ψ*

2. Two-Segment Model

For the model with two breakpoints (with initial guesses at *median*(*t_year_*) − 2 and *median*(*t_year_*) + 2, the regression is:s

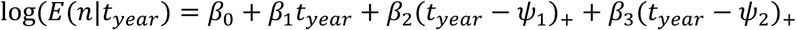

where:

- *Ψ*_1_ and *Ψ*_2_are estimated breakpoint.
- (*t_year_* − *Ψ*)_+_ is defined similarly for *i* = 1, 2

3. Three-Segment Model

For the model with three breakpoints (with initial guesses at *median*(*t_year_*) − 3, and *median*(*t_year_*) + 3, the regression is:

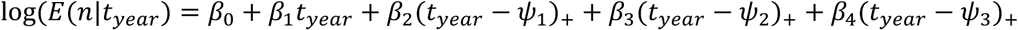

where:

- *Ψ*_1_, *Ψ*_2_ and *Ψ*_3_are estimated breakpoint.

Model fits with varying increasing number of segments, evaluated using AIC (lower values indicate better performance)

**S6 Figure 2.**
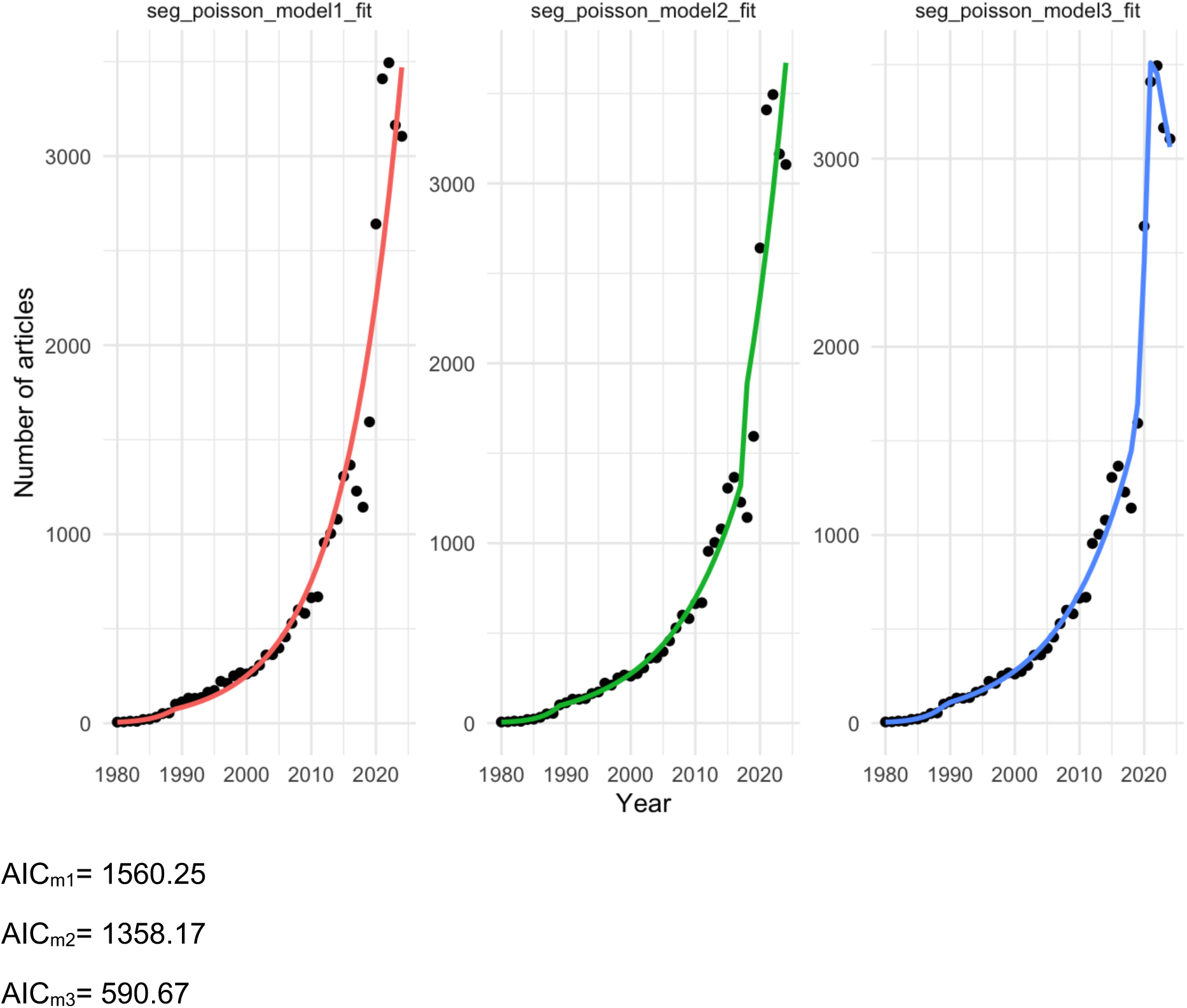

### S7 Results

**Figure 3.**
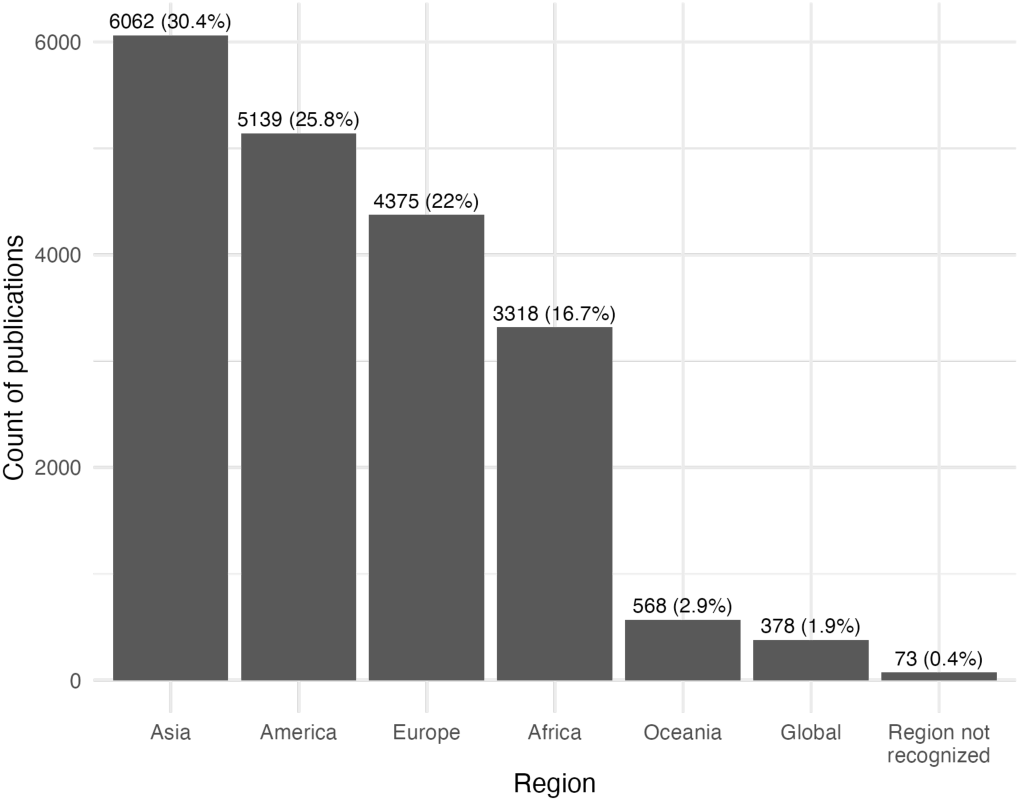
Distribution of geography-specific infectious disease modelling literature published between 1980 and 2024.

**S8 Table 3.** Countries of which policy documents identified by Overton to be publishing infectious disease modelling (IDM) literature by World Bank Economic Classification. Abbreviations: OECD = Organisation for Economic Co-operation and Development; H = high income country; USA = United States of America; UK = United Kingdom.

| **World Bank**<br>**Country**<br>**Classification**<br>**(2019)** | **Country** | **OECD country** | **Count of country** |
| --- | --- | --- | --- |
| **High income** | USA | x | 342 |
|  | UK | x | 83 |
|  | Canada | x | 66 |
|  | Spain | x | 51 |
|  | France | x | 47 |
|  | Germany | x | 47 |
|  | Switzerland | x | 34 |
|  | Netherlands | x | 23 |
|  | Australia | x | 23 |
|  | Japan | x | 15 |
|  | Ireland | x | 14 |
|  | Sweden | x | 13 |
|  | Finland | x | 11 |
|  | Norway | x | 11 |
|  | Belgium | x | 10 |
|  | New Zealand | x | 10 |
|  | Austria | x | 7 |
|  | Italy | x | 6 |
|  | Portugal | x | 5 |
|  | Estonia | x | 3 |
|  | Denmark | x | 2 |
|  | Andorra |  | 2 |
|  | Iceland | x | 2 |
|  | Taiwan |  | 1 |
|  | Luxembourg | x | 1 |
|  | Singapore |  | 1 |
|  | Uruguay |  | 1 |
|  | Slovenia | x | 1 |
|  | Hungary | x | 1 |
| **Upper middle**<br>**income** | South Africa |  | 14 |
|  | Brazil |  | 9 |
|  | Peru |  | 5 |
|  | Indonesia |  | 3 |
|  | Malaysia |  | 2 |
|  | Argentina |  | 2 |
|  | Thailand |  | 2 |
|  | Ecuador |  | 1 |
|  | Serbia |  | 1 |
|  | Colombia |  | 1 |
| **Lower middle**<br>**income** | Kenya |  | 3 |
|  | Philippines |  | 2 |
|  | India |  | 2 |
|  | Tunisia |  | 1 |
|  | Egypt |  | 1 |
|  | Sri Lanka |  | 1 |
|  | Cape Verde |  | 1 |
| **Low income** | Uganda |  | 5 |
|  | South Korea |  | 1 |

## Notes

### Competing Interest Statement

The authors have declared no competing interest.

### Funding Statement

This study did not receive any funding.

